# Prevalence of Elevated NT-proBNP and its Prognostic Value by Blood Pressure Treatment and Control- National Health and Nutrition Examination Survey, 1999-2004

**DOI:** 10.1101/2023.02.20.23286211

**Authors:** Natalie R. Daya, John W. McEvoy, Robert Christenson, Olive Tang, Kathryn Foti, Stephen P. Juraschek, Elizabeth Selvin, Justin B. Echouffo Tcheugui

## Abstract

**Background:** The prognostic utility of NT-proBNP in the setting of hypertension has not been well-characterized in the general US adult population.

**Methods:** We measured NT-proBNP among adults aged 20 years who participated in the 1999-2004 National Health and Nutrition Examination Survey. In adults without a history of cardiovascular disease, we assessed the prevalence of elevated NT-pro-BNP by blood pressure (BP) treatment and control categories. We examined the extent to which NT-proBNP identifies participants at higher risk for mortality across BP treatment and control categories.

**Results:** The number of US adults without CVD with elevated NT-proBNP (≥125 pg/ml) was 6.2 million among those with untreated hypertension, 4.6 million among those with treated controlled hypertension, and 5.4 million among those with treated uncontrolled hypertension. After adjusting for age, sex, body mass index, and race/ethnicity, participants with treated controlled hypertension and elevated NT-proBNP had increased risk of all-cause mortality (HR 2.29, 95% CI 1.79, 2.95) and increased risk of cardiovascular mortality (HR 3.83, 95% CI: 2.34, 6.29), compared to those without hypertension and with low levels of NT-proBNP (<125 pg/ml). Among those on antihypertensive medication, those with SBP 130-139 mm Hg and elevated NT-proBNP had increased risk of all-cause mortality, compared to those with SBP<120 mm Hg and low levels of NT-proBNP.

**Conclusions:** Among a general population of adults free of cardiovascular disease, NT-proBNP can provide additional prognostic information within and across categories of BP. Measurement of NT-proBNP may have potential for clinical use to optimize hypertension treatment.

## INTRODUCTION

N-terminal pro-B-type natriuretic peptide (NT-proBNP), a stable amino acid co-secreted with brain natriuretic peptide, is synthesized by the heart in reaction to cardiac wall stress. In physiological studies, natriuretic peptides maintain salt and water homeostasis, and regulate blood pressure (BP) through vasodilation, natriuresis, and increased urine output.^1^ In population-based studies, NT-pro-BNP has been shown to be associated with hypertension development.^2-5^ While NT-proBNP is a well-established marker of elevated cardiovascular disease risk and mortality,^6^ few studies have quantified how elevated NT-proBNP may help identify persons at risk for morbidity and mortality across the spectrum of BP treatment and control categories.^7,8^ Knowledge of the prognostic value of NT-proBNP in the setting of hypertension may better inform our approach to BP management. NT-proBNP can signal end-organ damage and may help guide intensification of BP therapy or provide additional value when BP levels relatively low signaling the need to initiate hypertension treatment. However, the potential impact of a biomarker augmented approach to BP management has not been assessed in large representative US population.

We measured NT-proBNP in stored samples from the 1999-2004 National Health and Nutrition Examination (NHANES) and assessed the prevalence of elevated NT-pro-BNP by BP treatment and control in this nationally representative sample of US adults 20 years of age and older. We also examined the degree to which NT-proBNP helped identify US adults at high risk for all-cause and cardiovascular mortality across BP categories; overall and according to subgroups defined by race/ethnicity and body mass index.

## METHODS

### Study Population

The NHANES is a large, nationally representative cross-sectional survey of the non-institutionalized civilian population in the US. We measured NT-proBNP in all participants who underwent a blood draw in the mobile examination center over the course of 3 combined survey cycles (1999-2000, 2001-2002 and 2003-2004) and had blood stored for future studies. Of the 14,213 eligible participants aged 20 years and older, we excluded participants missing specimens (n=1,903), missing data on mortality linkage (n=12), missing systolic or diastolic BP (n=532), as well as those with self-reported prevalent cardiovascular disease (n=1,384) defined as having been previously told by a physician or other health professional that they had congestive heart failure, coronary heart disease, angina, heart attack, or stroke. Our analytic sample included 10,382 participants. The NHANES protocols and this stored serum study was approved by the ethics review board of the National Center for Health Statistics. All participants provided written informed consent.

### N-terminal pro-B-type natriuretic peptide (NT-proBNP)

We measured NT-proBNP in all available surplus serum samples between 2018-2020 at the University of Maryland School of Medicine (Baltimore, Maryland, USA). NT-proBNP was measured using the Roche e611 (Roche Diagnostics Corp) autoanalyzer. The lower and upper limits of detection were 5 pg/ml and 35,000 pg/ml, respectively. The coefficient of variations for NT-proBNP were 3.1% (low, 46 pg/ml) and 2.7% (high, 32805 pg/ml).

We primarily defined elevated NT-proBNP as the clinical cut point of ≥125 pg/ml, but also examined the following cut-points: ≥300 pg/ml and ≥450 pg/ml. For assessing the mortality outcomes, we used the following categories: <125 pg/ml, ≥125 pg/ml; <300 pg/ml, ≥300 pg/ml; and <450 pg/ml, ≥450 pg/ml.

### BP Treatment and Control Categories

Trained physicians obtained BP measurements during the NHANES examination using a standard study protocol.^9^ Three consecutive auscultatory BP readings were obtained using a mercury sphygmomanometer and appropriate-size BP cuff after the participant rested in a seated position for 5 minutes. If any of the BP readings were interrupted or incomplete, a fourth attempt was made. All available readings were used to calculate mean systolic BP (SBP) and diastolic BP (DBP). Antihypertensive medication use was self-reported.

BP treatment and control categories were defined as (1) No hypertension (no self-reported use of antihypertensive medication and SBP<140 mm Hg and DBP<90 mm Hg); (2) Untreated hypertension (no self-reported use of antihypertensive medication and SBP≥140 mm Hg or DBP≥90 mm Hg); (3) Treated controlled hypertension (self-reported use of antihypertensive medication and SBP<140 mm Hg and DBP<90 mm Hg); and (4) Treated uncontrolled hypertension (self-reported use of antihypertensive medication and SBP≥140 mm Hg or DBP≥90 mm Hg).

Alternatively, we stratified by current use of antihypertensive medication and categorized by BP levels – SBP was categorized as: <120, 120 to 129, 130 to 139, 140 to149 and ≥150 mm Hg; and DBP as: <60, 60 to 69, 70 to 79, 80 to 89 and ≥90 mm Hg.

### Mortality

Vital status was determined through a probabilistic match between NHANES personal identifiers and linkage to death certificates from the National Death Index through December 31, 2019.^10^ Cardiovascular mortality was identified based on the underlying cause of death from International Classification of Diseases (ICD) codes I00-I99.

### Covariates

A computer-assisted personal interview system was used to collect information on participants ‘ sociodemographic and lifestyle characteristics, including race/ethnicity, education (less than high school, high school diploma, more than high school), smoking status (current, former, never), alcohol consumption [current heavy (male>2 drinks/day, female>1 drink/day), current moderate (male ≤2 drinks/day, female ≤1 drink/day), former, never] and physical activity (at least 10 minutes of moderate or vigorous activity over the past 30 days).

We calculated body mass index (BMI) using height and weight measured at the examination. Diagnosed diabetes was defined as a self-reported physician diagnosis of diabetes or use of diabetes medication. Total cholesterol was measured using an enzymatic method. Hypercholesterolemia was defined as total cholesterol ≥240 mg/dL or self-reported lipid-lowering medication use. Estimated glomerular filtration rate (eGFR) was calculated using the 2021 CKD-EPI creatinine-cystatin C equation.^11^

### Statistical Analysis

All analyses were weighted and accounted for the complex sample survey design to generate estimates generalizable to the 1999-2004 US adult population. We summarized the baseline characteristics (means and percentages with corresponding standard errors using Taylor series linearization) by BP treatment and control categories. We presented the prevalence of elevated NT-proBNP (≥125 pg/ml, ≥300 pg/ml or ≥450 pg/ml) across BP treatment and control categories in millions using the 2003-2004 U.S. Census Population.

We examined the cross-sectional associations of BP treatment and control with elevated NT-proBNP (defined as ≥125 pg/ml, ≥300 pg/ml, or ≥450 pg/ml) using logistic regression. We conducted prospective analyses of the joint associations of BP treatment and control categories and NT-proBNP, using the commonly used clinical cut point of 125 mg/dl, with all-cause and cardiovascular mortality using Cox proportional hazards models. Model 1 included age, sex, and race/ethnicity. Model 2 included all variables in Model 1 plus education, alcohol use, smoking status, physical activity, BMI, diabetes, hypercholesterolemia, and eGFR.

For the analyses of the joint association of SBP categories and NT-proBNP with all-cause and cardiovascular mortality, the reference group was SBP <120 mm Hg and NT-proBNP<125 mg/dl. For the analyses of the joint association of DBP and NT-proBNP with all-cause and cardiovascular mortality, the reference group was DBP 70-79 mm Hg and NT-proBNP<125 mg/dl. We adjusted for variables in model 2 and stratified by current use of antihypertensive medication.

We conducted a sensitivity analysis using an alternate categorization of BP control, BP greater than or equal to 130/80 mm Hg, which is the current threshold of hypertension according to the 2017 American College of Cardiology/American Heart Association guidelines.^12^ We conducted sensitivity analyses in subgroups defined by race/ethnicity and by BMI categories (underweight: <18.5 kg/m^2^, normal weight: 18.5-<25 kg/m^2^, overweight: 25-<30 kg/m^2^, and obese: ≥30 kg/m^2^). We examined the joint association of BP treatment and control categories and NT-proBNP with all-cause and cardiovascular mortality. We tested for interaction by race and BMI, as these variables have been shown to affect levels of NT-proBNP.^13-15^ We also conducted sensitivity analyses using an alternate categorization of NT-proBNP (<300 pg/ml, ≥300 pg/ml and <450 pg/ml, ≥450 pg/ml)

Stata version 17.0 (College Station, Texas, USA) was used for all analyses. A two-sided *P*-value<0.05 was considered statistically significant.

## RESULTS

### Characteristics of Study Sample

Our study population included 10,382 participants (mean age 44.3 years; 52.3% women, 71.5% non-Hispanic White), of whom 3,270 (31.5%) had treated or untreated hypertension. Those with hypertension were more likely to be older, to be non-Hispanic Black adults, adults with obesity, current moderate or former drinkers, former smokers, have hypocholesterolemia, lower eGFR, higher levels of NT-proBNP, and report lower levels of physical activity. Individuals with untreated hypertension tended to be younger, have a lower BMI, and less likely to have diabetes or hypocholesterolemia, than those with treated (controlled or uncontrolled) hypertension (**Table 1**).

**Table 1.** **Characteristics of US adults according to blood pressure treatment and control categories, US adults aged 20 years or older, NHANES 1999-2004 (N=10,382)**.

|  | No<br>hypertension | Untreated<br>hypertension | Treated<br>controlled<br>hypertension | Treated<br>uncontrolled<br>hypertension |
| --- | --- | --- | --- | --- |
| **Unweighted N** | 7112 | 1386 | 1021 | 863 |
| **Age, years (mean (SE))** | 39.7 (0.3) | 55.2 (0.8) | 56.7 (0.5) | 62.5 (0.6) |
| **Age categories, % (SE)** |  |  |  |  |
| 20-39 years | 54.1 (1.0) | 18.1 (2.1) | 8.4 (0.9) | 5.5 (1.1) |
| 40-64 years | 40.3 (1.0) | 51.0 (1.8) | 62.0 (1.6) | 46.9 (1.9) |
| 65-79 years | 4.8 (0.3) | 23.1 (1.4) | 25.0 (1.6) | 35.8 (1.9) |
| ≥80 years | 0.8 (0.1) | 7.9 (0.8) | 4.6 (0.7) | 11.8 (1.2) |
| **Male, % (SE)** | 48.5 (0.6) | 51.1 (1.7) | 44.1 (1.8) | 37.3 (2.4) |
| **Race/Ethnicity, % (SE)** |  |  |  |  |
| Non-Hispanic White | 70.9 (1.6) | 71.7 (2.5) | 76.7 (2.2) | 70.8 (2.8) |
| Non-Hispanic Black | 9.6 (0.8) | 12.4 (1.6) | 12.7 (1.5) | 16.0 (2.0) |
| Mexican American | 8.7 (0.9) | 6.1 (1.1) | 3.0 (0.8) | 3.9 (1.0) |
| Other/Other Hispanic | 10.7 (1.4) | 9.8 (1.9) | 7.5 (1.5) | 9.3 (1.6) |
| **Systolic BP, mm Hg (mean (SE))** | 115.4 (0.2) | 149.6 (0.7) | 123.0 (0.4) | 156.3 (0.9) |
| **Diastolic BP, mm Hg (mean(SE))** | 70.2 (0.2) | 81.4 (0.7) | 71.3 (0.6) | 77.2 (0.9) |
| **BMI categories, % (SE)** |  |  |  |  |
| Underweight | 2.2 (0.2) | 1.9 (0.6) | 0.5 (0.3) | 0.2 (0.1) |
| Normal | 38.0 (0.8) | 23.9 (1.5) | 14.5 (1.5) | 19.6 (1.8) |
| Overweight | 34.5 (1.0) | 38.3 (1.8) | 34.6 (1.6) | 32.6 (2.1) |
| Obese | 25.3 (0.8) | 35.9 (2.0) | 50.3 (1.7) | 47.6 (2.1) |
| **Diabetes, % (SE)** | 3.1 (0.3) | 5.3 (0.8) | 15.7 (1.1) | 16.4 (1.2) |
| **Alcohol use, % (SE)** |  |  |  |  |
| Current moderate | 33.7 (1.3) | 34.2 (2.0) | 37.2 (2.1) | 37.6 (2.0) |
| Current heavy | 41.6 (1.1) | 31.2 (1.8) | 25.6 (2.2) | 19.3 (2.2) |
| Former | 13.5 (0.7) | 20.3 (1.5) | 22.7 (1.7) | 23.5 (2.0) |
| Never | 11.1 (1.2) | 14.3 (1.7) | 14.5 (1.7) | 19.6 (1.8) |
| **Smoking status, % (SE)** |  |  |  |  |
| Current | 27.9 (1.0) | 19.2 (1.3) | 17.8 (1.4) | 12.0 (1.3) |
| Former | 20.6 (0.9) | 30.8 (1.6) | 35.6 (2.1) | 30.8 (2.2) |
| Never | 51.5 (1.3) | 50.0 (1.5) | 46.6 (1.8) | 57.2 (2.2) |
| **Physical activity (moderate to vigorous), % (SE)** | 69.1 (1.1) | 59.3 (1.8) | 60.3 (2.0) | 55.3 (1.9) |
| **Hypercholesterolemia, % (SE)** | 17.9 (0.7) | 31.9 (1.5) | 42.5 (2.2) | 42.3 (2.1) |
| **eGFR-CrCys (2021), mean** | 128.9 (0.4) | 120.2 (0.7) | 114.4 (0.8) | 110.8 (1.1) |
| (SE) |  |  |  |  |
| **NT-proBNP, pg/ml** |  | 59.3 (26.1, | 57.0 (28.8, | 101.6 (54.2, |
| **(median (IQR))** | 37.2 (19.2, 69.6) | 143.1) | 124.8) | 215.2) |
| **NT-proBNP ≥125 pg/ml,** |  |  |  |  |
| **% (SE)** | 9.1 (0.4) | 27.2 (1.4) | 24.9 (1.5) | 43.3 (2.2) |
| **NT-proBNP ≥300 pg/ml,** |  |  |  |  |
| **% (SE)** | 1.6 (0.2) | 9.2 (1.0) | 8.6 (0.9) | 18.6 (1.6) |
| **NT-proBNP ≥450 pg/ml,** |  |  |  |  |
| **% (SE)** | 0.8 (0.1) | 5.3 (0.8) | 6.2 (0.6) | 11.1 (1.2) |

IQR: interquartile range
No hypertension: no self-reported use of antihypertensive medication and SBP<140 mm Hg and DBP<90 mm Hg
Untreated hypertension: no self-reported use of antihypertensive medication and SBP ≥ 140 mm Hg or DBP ≥ 90 mm Hg
Treated controlled hypertension: self-reported use of antihypertensive medication and SBP<140 mm Hg and DBP<90 mm Hg
Treated uncontrolled hypertension: self-reported use of antihypertensive medication and SBP ≥ 140 mm Hg or DBP ≥ 90 mm Hg

### Prevalence of elevated NT-proBNP according to categories of BP treatment and control

The prevalence of elevated NT-proBNP (≥125 pg/ml) among US adults with untreated hypertension was 27.2% (95% CI 24.4-30.2), representing approximately 6.2 million (95% CI 5.0-7.6) adults (**Figure 1; Table S1**). The prevalence of elevated NT-proBNP among those with treated controlled hypertension was 24.9% (95% CI 22.1-28.0), representing approximately 4.6 million (95% CI 3.7-5.7) adults. The prevalence of elevated NT-proBNP among those with treated uncontrolled hypertension was 43.3% (95% 38.9-47.8), representing approximately 5.4 million (95% CI 4.4-6.5) adults. The prevalence of NT-proBNP ≥300 pg/ml, ≥450 pg/ml (**Table S1**), and by SBP and DBP categories (**Tables S2 and S3**) are provided in the Supplement. Using these alternative cut-points for defining elevated NT-proBNP (≥300 pg/ml or ≥450 pg/ml), the highest prevalence estimates were observed among those with treated uncontrolled hypertension, or those with a SBP≥150 mmHg, or those with a DBP in the 70-79 mm Hg range.

**Figure 1.**
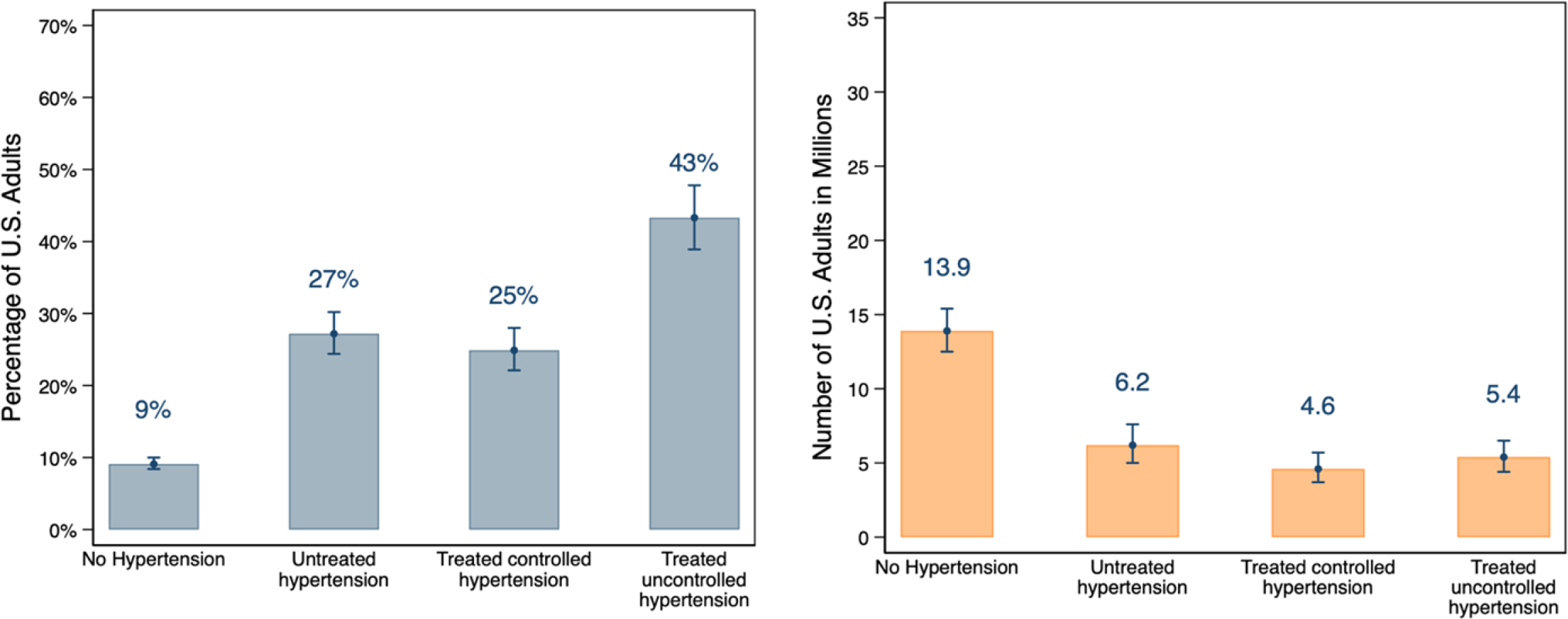
Prevalence of elevated NT-proBNP (≥125 pg/ml) among US adults aged 20 and older, by BP treatment and control categories, using the 2003-2004 U.S. census population.

Individuals with untreated hypertension or treated uncontrolled hypertension were more likely to have elevated NT-proBNP, compared to those without hypertension, even after adjustment for lifestyle and clinical risk factors (**Table 2**).

**Table 2.** **Cross-sectional associations of blood pressure treatment and control categories with elevated NT-proBNP**.

|  | No<br>hypertension | Untreated<br>hypertension | Treated<br>controlled<br>hypertension | Treated<br>uncontrolled<br>hypertension |
| --- | --- | --- | --- | --- |
| **Elevated NT-proBNP** | **OR (95% CI)** | **OR (95% CI)** | **OR (95% CI)** | **OR (95% CI)** |
| **NT-proBNP $\geq 125$ pg/ml** |  |  |  |  |
| Unweighted n/N | 725/7112 | 472/1386 | 294/1021 | 421/863 |
| Model 1 | 1 (reference) | 1.32 (1.11, 1.57) | 1.10 (0.88, 1.37) | 1.82 (1.48, 2.25) |
| Model 2 | 1 (reference) | 1.43 (1.20, 1.79) | 1.16 (0.88, 1.52) | 2.03 (1.57, 2.62) |
| **NT-proBNP $\geq 300$ pg/ml** |  |  |  |  |
| Unweighted n/N | 160/7112 | 178/1386 | 105/1021 | 199/863 |
| Model 1 | 1 (reference) | 1.50 (1.16, 1.95) | 1.56 (1.02, 2.39) | 2.44 (1.85, 3.21) |
| Model 2 | 1 (reference) | 1.53 (1.09, 2.14) | 1.47 (0.92, 2.34) | 2.72 (1.93, 3.85) |
| **NT-proBNP $\geq 450$ pg/ml** |  |  |  |  |
| Unweighted n/N | 92/7112 | 101/1386 | 72/1021 | 123/863 |
| Model 1 | 1 (reference) | 1.52 (1.03, 2.24) | 2.14 (1.35, 3.41) | 2.42 (1.55, 3.78) |
| Model 2 | 1 (reference) | 1.61 (0.99, 2.60) | 2.28 (1.35, 3.87) | 2.84 (1.54, 5.22) |

**Model 1:** age, sex, race/ethnicity
**Model 2:** all variables in Model 1 + education, alcohol use, smoking status, physical activity, BMI, diabetes, hypercholesterolemia, and eGFR.

Individuals with SBP≥150 mm Hg were more likely to have elevated NT-proBNP (irrespective of its definition) compared to those with SBP<120 mm Hg, regardless of antihypertensive medication use (**Table S4a and S4b**). Those with DBP levels <60 or 60-69 mm Hg and not using antihypertensive medication were more likely to have an NT-proBNP ≥125 pg/ml compared to those with DBP 70-79 mm Hg (**Table S5a**).

### Prospective associations of NT-proBNP and BP treatment and control with mortality

Over a median follow-up of 17.3 years, 2,369 participants died from all-causes, including 661 cardiovascular-related deaths. The crude all-cause and cardiovascular specific mortality rates across all levels of NT-proBNP were similar for those with untreated hypertension and treated controlled hypertension, lower for those with no hypertension, and highest for those with treated uncontrolled hypertension (**Table 3**). The crude all-cause mortality rates increased as NT-proBNP increased (**Figure 2**).

**Table 3.** **Mortality rates (per 1,000 person-years) (95% CI) and adjusted* HR (95% CI) of all-cause and cardiovascular specific mortality across categories of NT-proBNP, by blood pressure treatment and control categories**.

|  | No hypertension | Untreated hypertension | Treated controlled hypertension | Treated uncontrolled hypertension |
| --- | --- | --- | --- | --- |
|  | Mortality rate (95% CI) | Mortality rate (95% CI) | Mortality rate (95% CI) | Mortality rate (95% CI) |
| **All-cause mortality** |  |  |  |  |
| **Overall** | 5.33<br>(4.92, 5.79) | 21.7<br>(19.6, 24.1) | 22.1<br>(19.6, 25.0) | 30.5<br>(27.2, 34.3) |
| **NT-proBNP categories** | **HR (95% CI)** | **HR (95% CI)** | **HR (95% CI)** | **HR (95% CI)** |
| <125 pg/ml | 1 (reference) | 1.44 (1.20, 1.73) | 1.23 (1.02, 1.49) | 1.42 (1.12, 1.80) |
| ≥125 pg/ml | 1.56 (1.26, 1.91) | 2.02 (1.66, 2.45) | 2.29 (1.79, 2.95) | 1.88 (1.46, 2.42) |
| **Cardiovascular mortality** | **Mortality rate (95% CI)** | **Mortality rate (95% CI)** | **Mortality rate (95% CI)** | **Mortality rate (95% CI)** |
| **Overall** | 1.08<br>(0.91, 1.29) | 5.26<br>(4.34, 6.42) | 6.58<br>(5.32, 8.23) | 11.7<br>(9.71, 14.2) |
| **NT-proBNP categories** | **HR (95% CI)** | **HR (95% CI)** | **HR (95% CI)** | **HR (95% CI)** |
| <125 pg/ml | 1 (reference) | 1.46 (0.99, 2.15) | 1.26 (0.88, 1.82) | 2.36 (1.60, 3.48) |
| ≥125 pg/ml | 2.13 (1.39, 3.26) | 2.58 (1.85, 3.60) | 3.83 (2.34, 6.29) | 3.60 (2.45, 5.27) |

\*Adjusted for age, sex, race/ethnicity, education, alcohol use, smoking status, physical activity, BMI, diabetes, hypercholesterolemia, and eGFR.

**Figure 2.**
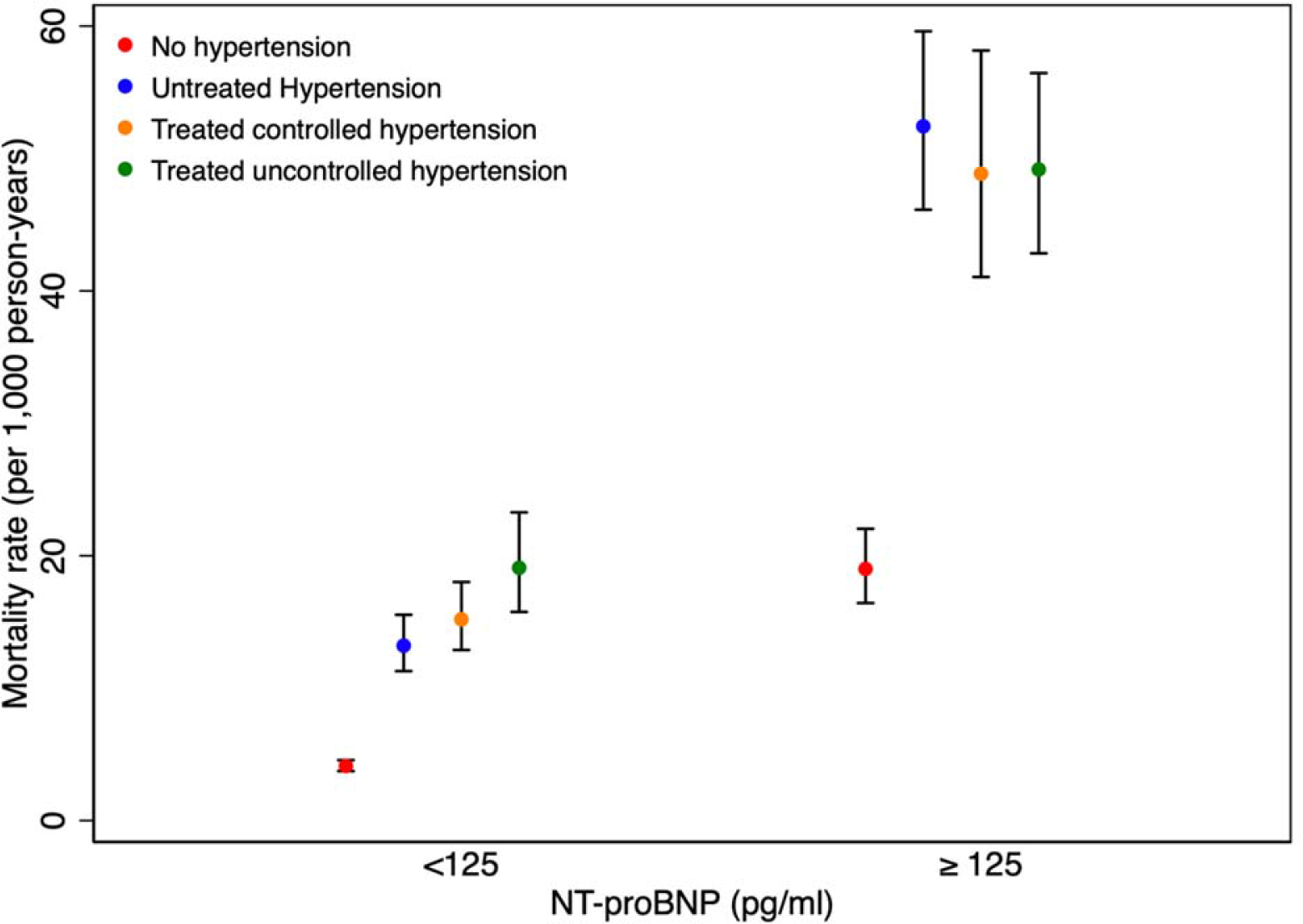
All-cause mortality rates (95% CI) across categories of BP treatment and control, and NT-proBNP.

In multivariable adjusted models, participants with treated controlled hypertension and NT-proBNP ≥125 pg/ml, had the highest relative risk of all-cause mortality (hazard ratio [HR]: 2.29, 95% CI: 1.79, 2.95) and cardiovascular mortality (HR: 3.83, 95% CI: 2.34, 6.29), compared to those without hypertension and low levels of NT-proBNP (<125 pg/ml) (**Table 3**). Those with untreated hypertension and NT-proBNP ≥125 pg/ml had the second highest risk of all-cause mortality (HR: 2.02, 95% CI: 1.66, 2.45) and those with treated uncontrolled hypertension and NT-proBNP ≥125 pg/ml had the second highest risk of cardiovascular mortality (HR: 3.60, 95% CI: 2.45, 5.27). Similar patterns of BP treatment and control with mortality outcomes were observed when using alternative definitions of BP control (130/80 mm Hg) (**Table S6**) and elevated NT-proBNP (≥300 pg/ml or ≥450 pg/ml) (**Tables S7 and S8**).

Race/ethnicity and BMI did not significantly modify the association of quartiles of NT-proBNP and BP treatment and control categories with mortality (p for interaction>0.05, **Tables S9 and S10**).

### Associations of NT-proBNP, SBP or DBP levels with Mortality

Crude all-cause and cardiovascular mortality rates tended to increase across levels of SBP and among those on antihypertensive medication and decrease across levels of DBP **(Tables 4 and S11)**. The adjusted hazard ratio of all-cause and cardiovascular mortality generally increased as SBP or NT-proBNP increased among those not on antihypertensive medication. There was a U-shaped association between increasing levels of DBP and the risk of mortality, irrespective of antihypertensive medication use. Among participants not on antihypertensive medication, individuals with SBP≥150 mm Hg and elevated NT-proBNP had the highest risk of all-cause mortality and those with SBP 120-<130 mm Hg and elevated NT-proBNP had the highest risk of cardiovascular mortality (**Table 4a**). Regarding participants on antihypertensive treatment, those with SBP 130-<140 mm Hg and elevated NT-proBNP had the highest risk of all-cause and those with SBP≥150 mm Hg and elevated NT-proBNP had the highest risk of cardiovascular mortality (**Tables 4b**). Among participants on antihypertensive treatment, those with DBP <60 mmHg and elevated NT-proBNP had the highest risk of all-cause mortality and those with DBP ≥90 mm Hg had the highest risk of cardiovascular specific mortality (**Table S11 & S14**).

**Table 4.**
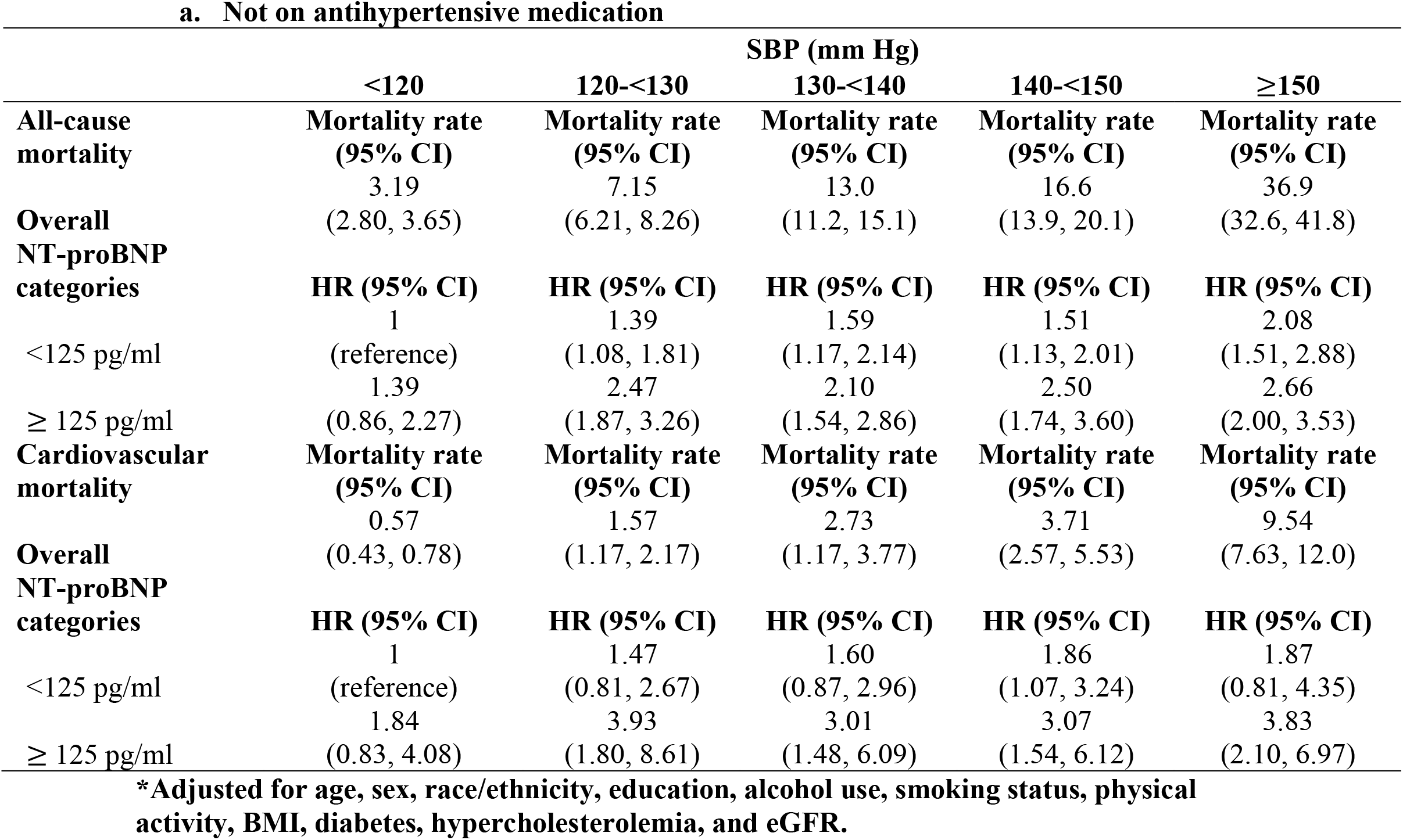
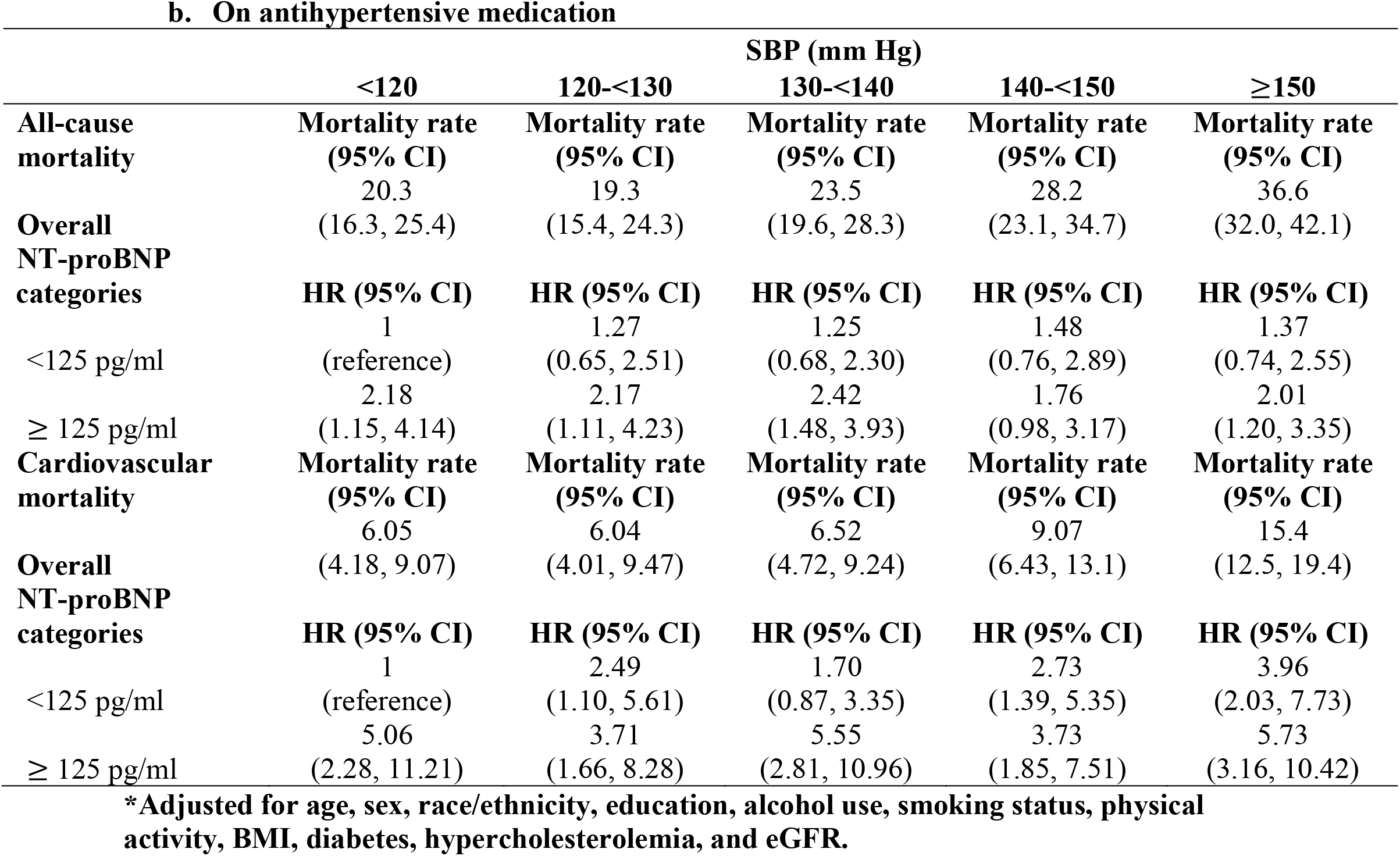
**Crude mortality rate (per 1,000 person-years) (95% CI) and adjusted* HR (95% CI) of all-cause and cardiovascular specific mortality across quartiles of NT-proBNP, by systolic blood pressure categories among those (a) not on antihypertensive medication and those (b) on antihypertensive medication**.

## DISCUSSION

Our study provides national prevalence data on the extent of elevated NT-proBNP across levels of BP. The number of individuals with elevated NT-proBNP (≥125 mg/dL) in the US population, was approximately 13.9 million adults, or 9%, among those with no hypertension, 6.2 million, or 27%, among those with untreated hypertension, 4.6 million, or 25%, among those with treated controlled hypertension, and 5.4 million, or 43%, among those with treated uncontrolled hypertension.

In this nationally representative sample of the US adult population, we observed that the risk of all-cause mortality was higher among individuals with treated controlled or untreated hypertension and elevated NT-proBNP than the corresponding risks among those without hypertension or with treated uncontrolled hypertension. Furthermore, individuals on antihypertensive medication and with SBP 130 to 139 mm Hg, DBP <60 mm Hg, or ≥90mmHg, and with elevated NT-proBNP, tended to have the highest risk of all-cause and/or cardiovascular mortality. Our findings suggest that NT-proBNP is associated with adverse outcomes across the BP/hypertension treatment spectrum, thus suggesting that an NT-proBNP augmented approach to BP management at the population level is potentially appropriate. This may include BP therapy intensification guided by levels of NT-proBNP.

While NT-proBNP is frequently used in patients suspected to have heart failure for diagnosis and monitoring of heart failure therapy,^16^ it is not currently used as a tool to refine or augment BP therapy. A prior study in the Atherosclerosis Risk in Communities (ARIC) Study showed that within categories of BP, higher NT-proBNP (100 to <300 or 300 pg/ml) was associated with a graded increased risk for cardiovascular disease events and mortality, as compared to lower NT-proBNP (<100 pg/ml).^7^ In the ARIC Study, participants with SBP 130 to 139 mm Hg and NT-proBNP ≥300 pg/ml had a 3.5-fold higher risk of cardiovascular disease, compared with an NT-proBNP of <100 pg/ml and SBP of 140 to 149 mm Hg.^7^ Our study complements and extends the results of this prior study by providing results from a sample that is representative of the general US population, including a more diverse population in terms of age, sex, race/ethnicity, and other characteristics.^7,8^ The representativeness of our sample is of importance given the persistently higher rates of uncontrolled BP among younger individuals (age 18 to 44 years) compared to older individuals (age 45 years and above) in the general US population.^17^

Our results support the American College of Cardiology/American Heart Association recommendation to individualize BP targets using background cardiovascular disease risk rather than solely relying on the absolute value of BP.^18^ NT-proBNP has been shown to be a robust marker of elevated cardiovascular disease risk,^19^ specifically among individuals not on BP lowering therapy.^8^ The routine use of NT-proBNP in hypertension patients could indicate end-stage organ damage, even among individuals already on therapy. End-stage organ damage includes individuals with malignant left ventricular hypertrophy, for example, who may need BP intensive BP lowering.^20^ It could also help trigger assessments for a secondary cause of hypertension (metabolic, renal or other causes) among those with uncontrolled BP despite optimal therapy (i.e. resistant hypertension).^21^ We observed elevated mortality among those with high NT-proBNP in lower categories of BP, suggesting that NTproBNP can “unmask” elevated risk in uncontrolled BP.^22,23^ Our results suggest that NT-proBNP provides insight into possible end-organ damage and thus the need for early initiation of hypertension treatment.

### Strengths and limitations

Our study has limitations. First, despite the large sample size, the number of deaths in some cross-categories of NT-proBNP and BP were small, which limited our ability to examine mortality in those subgroups. Second, our assessment of death and its causes was limited to the use of ICD codes, which may underestimate the rates of cardiovascular disease-specific death. Third, we were unable to assess the risk of nonfatal cardiovascular disease events. Although in prior studies, the link between NT-proBNP and mortality have generally paralleled its associations with nonfatal cardiovascular disease events.^19^ Finally, as our study is observational there is the possibility of residual confounding and that the observed findings may not reflect causal associations.

The strengths of this study include the ability to generalize our findings to the general adult US population. We conducted rigorous and extensive adjustments for relevant confounders and accounted for treated and untreated hypertension. Moreover, we were able to test the robustness of our findings by levels of SBP and DBP, and accounting for the extent of BP control, all-cause and cardiovascular specific mortality, and categories of NT-proBNP that are most clinically relevant.^19,24^

## CONCLUSION

In the US adult population free of cardiovascular disease, NT-proBNP can provide risk information that could influence BP management, by identifying adults at higher risk of mortality across the stages of hypertension as well as among those without hypertension. Our study suggests that elevated levels of NT-proBNP across the BP treatment and control, as well as across levels of SBP and DBP, is an indicator of mortality. Furthermore, those who are on antihypertensive medication with SBP 130 to 139 mm Hg have the highest risk of all-cause mortality and those who are not on antihypertensive medication with SBP 120 to 129 mm Hg have the highest risk of cardiovascular mortality. Such individuals might derive the greatest benefit from intensive optimization of their BP control. Future trials should consider validation of a biomarker-assisted approach to guide clinical decisions surrounding BP treatment initiation or titration.

## Data Availability

The datasets generated and analyzed during the current study are available from the corresponding author, Natalie R Daya, on reasonable request. All NHANES data is publicly available.

https://wwwn.cdc.gov/nchs/nhanes/Default.aspx

## ACKNOWLEDGEMENTS

Reagents for the NT-proBNP assay were donated by the Roche Diagnostics Corporation.

## SUPPLEMENTAL MATERIAL

Tables S1-S15

